# The effectiveness and safety of acupuncture kinesitherapy after percutaneous coronary intervention in patients with coronary heart disease: study protocol for a randomized controlled trial

**DOI:** 10.1101/2022.07.10.22277473

**Authors:** Di Zhang, Huaimin Lu, Song Jin, Hong Guo, Xiaoxiao Liu, Wensheng Xiao, Hongpeng Li, Jiang Ma

## Abstract

**Background:** Coronary heart disease(CHD) is a common disease of the cardiovascular system. Percutaneous coronary intervention(PCI) has been proven an effective treatment. Various complications after PCI may further affect the long-term efficacy of PCI. Previous trials have showed that both kinesitherapy(KT) and acupuncture are beneficial to patients with CHD, but need further confirmation. Acupuncture kinesitherapy(AKT) is a kind of combination therapy of acupuncture with exercise training. In this study, we plan to conduct a randomized controlled trial(RCT) to confirm the superior efficacy of AKT compared to KT and conventional medical therapy(MT) in post-PCI rehabilitation for CHD patients, and to further guide the clinical strategy.

**Methods:** This is a single-center randomized controlled trial with 3 parallel arms. We will recruit 102 CHD patients after PCI and randomly assign them to 3 groups. Participants in 3 groups will receive 2-week MT. Besides, participants in AKT and KT groups will also receive acupuncture kinesitherapy and kinesitherapy alone, respectively. They will be treated once a day, 5-day a week, and the treatment course will last for a total of 2-week with a 2-day off during the course. The primary outcome is the changes in cardiac function. The secondary outcomes include aerobic capacity, muscle strength, flexibility, balance, coordination, mental status, and activity of daily living. All outcomes will be measured at baseline and 2-week after treatment. Adverse events also will be recorded. The cardiac function and activities of daily living will be followed up at 1, 3 and 6 months after treatment.

**Discussion:** We expect findings of trial will provide important insight into application of AKT as a safety and more effective method for CHD patients. Successful completion of this proposed study will also contribute to promotion of AKT in the clinic in the future.

**Trial registration:** Chinese clinical trial registry (Register Number: ChiCTR2100048960) http://www.chictr.org.cn/index.aspx.

## 1. Introduction

Coronary heart disease (CHD), also known as coronary artery disease (CAD), is a common disease of the cardiovascular system, and also the main cause of death in patients with cardiovascular disease [1]. The number of patients with cardiovascular diseases in China reached 330 million, among which the number of CHD was 11 million, according to the summary of the China Cardiovascular Health and Disease Report 2019 [2]. Percutaneous coronary intervention (PCI), a transcardiac catheter technology which dredges stenosis or occlusion of the coronary artery lumen to improve the blood perfusion of the myocardium, has been proven an effective treatment to reduce mortality and morbidity in patients of CHD [3-6]. However, PCI cannot reverse or slow down the process of coronary atherosclerosis and eliminate the risk factor for CHD. Coronary artery stenosis or occlusion results in myocardial ischemia, necrosis, and cardiac dysfunction, which further follows by various complications after PCI, including acute coronary artery occlusion, coronary spasm, coronary artery perforation, and even restenosis or thrombus, which may further affect the long-term efficacy of PCI [7-9]. Medical therapy (MT) including tirofiban, calcium antagonist and aspirin is usually applied for the treatment of complications after PCI. However, the long-term medication accompanies with inevitable gastrointestinal side effects such as bleeding, nausea, vomiting, etc., and cannot continuously and effectively improve the prognosis after PCI [10-12].

Kinesitherapy (KT), a central element of cardiac rehabilitation programs, is recognized as integral to the comprehensive care and has been given a Class I recommendation from the American Heart Association, the American College of Cardiology, and the European Society of Cardiology [3,12-14]. Researches on the efficacy of KT for CHD patients after PCI are inconsistent. Several studies [15-20] and systematic reviews [21] showed that KT can effectively improve cardiac and vascular functions for patients after PCI, and reduce the morbidity of cardiovascular events and the recurrence and mortality of CHD. The muscle strength, muscle endurance, balance, coordination ability, and quality of life have also been improved by KT, as well as the safety of KT [22-24]. However, two systematic reviews [25,26] showed that KT had no significant effect on restenosis, recurrence of myocardial infarction, improvement of cardiac function, and reduction of adverse cardiovascular events in patients after PCI.

Acupuncture kinesitherapy (AKT) is a kind of moving acupuncture therapy with exercise training carried out following acupuncture. Ocular acupuncture, an acupuncture treatment manipulated on orbit with 8 areas and 13 acupoints, has been approved by the Standardization Administration of China in 2009 [27]. Doctor Jingshan Peng, the professor of Liaoning University of Traditional Chinese Medicine, abstracted and summarized the theory of ocular acupuncture according to the Eight Profile and Meridian theories of the Traditional Chinese Medicine in the early 1970s. acupuncture around the orbital margin is recognized as a sort of micro-acupuncture, which can activate the meridians, invigorate blood, relieve pain and regulate viscera functions by stimulating acupoints around the orbital margin [28,29]. According to the Meridian Theory of the Traditional Chinese Medicine [30,31], the tissue around the eye is closely related to the Zang and Fu organs, and acupuncture around the eye is effective in treating heart diseases. Mao [32] found that acupuncture on the acupoints of the “Heart Area” and the “Upper Jiao Area” around the eye can significantly reduce the time and frequency of angina pectoris in patients with CHD, as well as the levels of TNF-α and CRP.

Studies show that the proportion of postoperative rehabilitation after PCI is about 60% in the United States, while it is still in its infancy in China [33,34]. Due to the frequent complications after PCI for CHD, it is imperative to carry out postoperative rehabilitation therapy after PCI in China. AKT may be an effective treatment for CHD patients after PCI. It is expected to optimize the post-PCI rehabilitation program by reducing complications after PCI, prolonging postoperative survival time, and improving the quality of life for CHD patients after PCI.

We plan to conduct a randomized controlled trial (RCT) to confirm the superior efficacy of AKT compared to KT and conventional medical therapy (MT) in post-PCI rehabilitation for patients of CHD. Changes in cardiac function, aerobic capacity, muscle strength, flexibility, balance, coordination, mental status, and activity of daily living will be measured to evaluate the efficacy of AKT comprehensively. Findings of this study are expected to provide an evidence-based decision on the application of AKT for CHD patients after PCI.

## 2. Methods and Design

### 2.1 Design

This is a single-center RCT with 3 parallel arms that aims to observe the effectiveness and safety of AKT for post-PCI rehabilitation and to compare the effects of AKT, KT, and MT. This trial has been approved by the Institutional Review Board of Hospital of Chengdu University of Traditional Chinese Medicine in July 2021 (reference number: 2021KL-028) and been registered (ChiCTR2100048960) at the Chinese Clinical Trial Registry in July 2021. The trial will be conducted in the Hospital of Chengdu University of Traditional Chinese Medicine. The whole study period is a 2-week intervention phase. After stratified randomization, participants in 3 groups will receive 2-week MT. Besides, participants in AKT and KT group will also receive acupuncture kinesitherapy and kinesitherapy alone, respectively. They will be treated once a day, 5 days a week, and the treatment course will last for a total of 2 weeks with a 2-day off during the course. Outcomes will be measured at baseline and 2 weeks after intervention, the cardiac function and activities of daily living will be followed up at 1, 3 and 6 months after treatment. All participants will be required to sign the written informed consent before randomization. The flow diagram of this trial is presented in **Fig. 1**. To provide a high quality of evidence, we will conduct this trial in line with Standard Protocol Items: Recommendations for Interventional Trials (SPIRIT) [35] and report our findings in accordance with the Consolidated Standards of Reporting Trial (CONSORT) statement [36].

**Fig 1.**
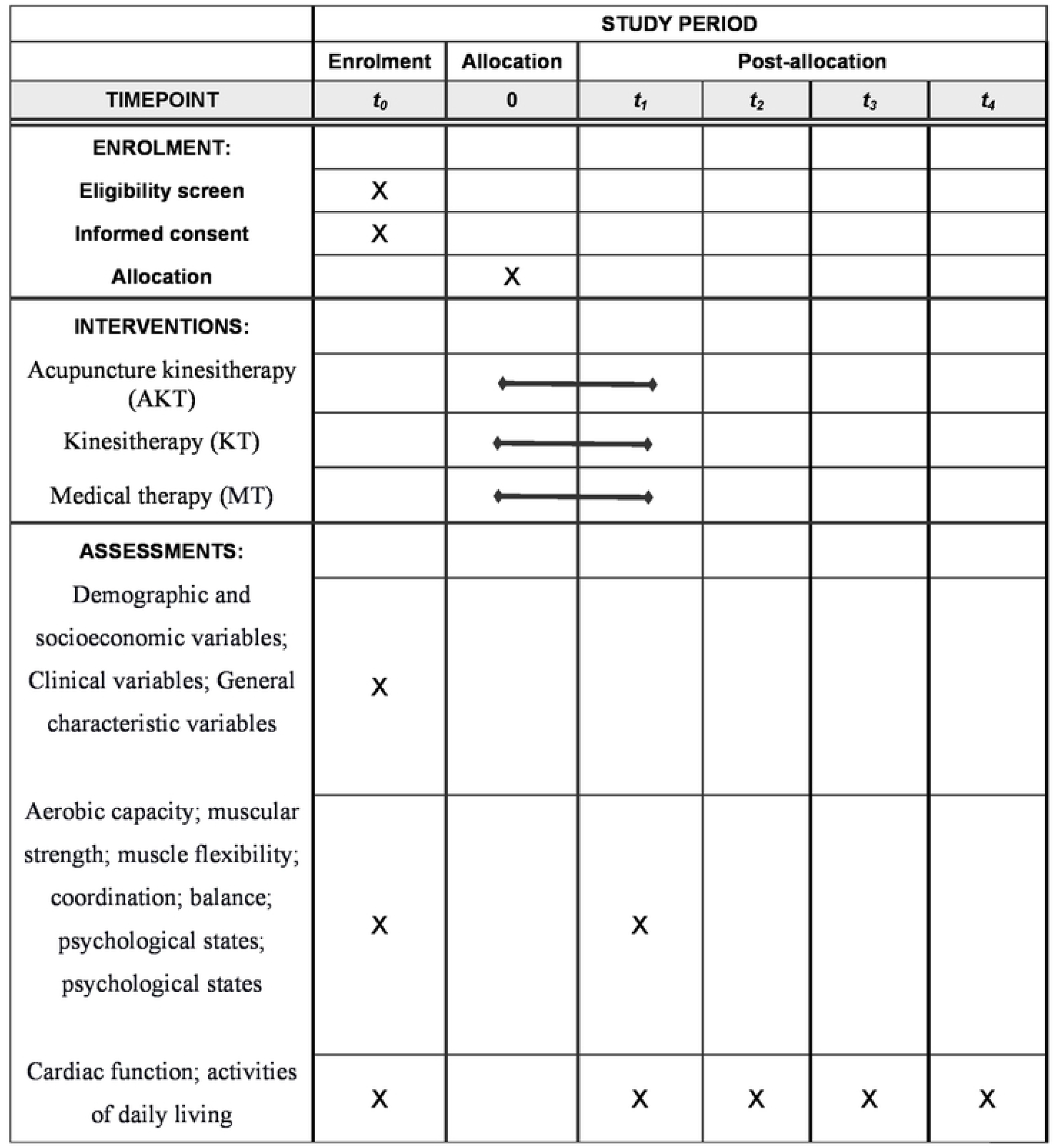
SPIRIT schedule of enrolment,interventions, and assessments.

**Fig 2.**
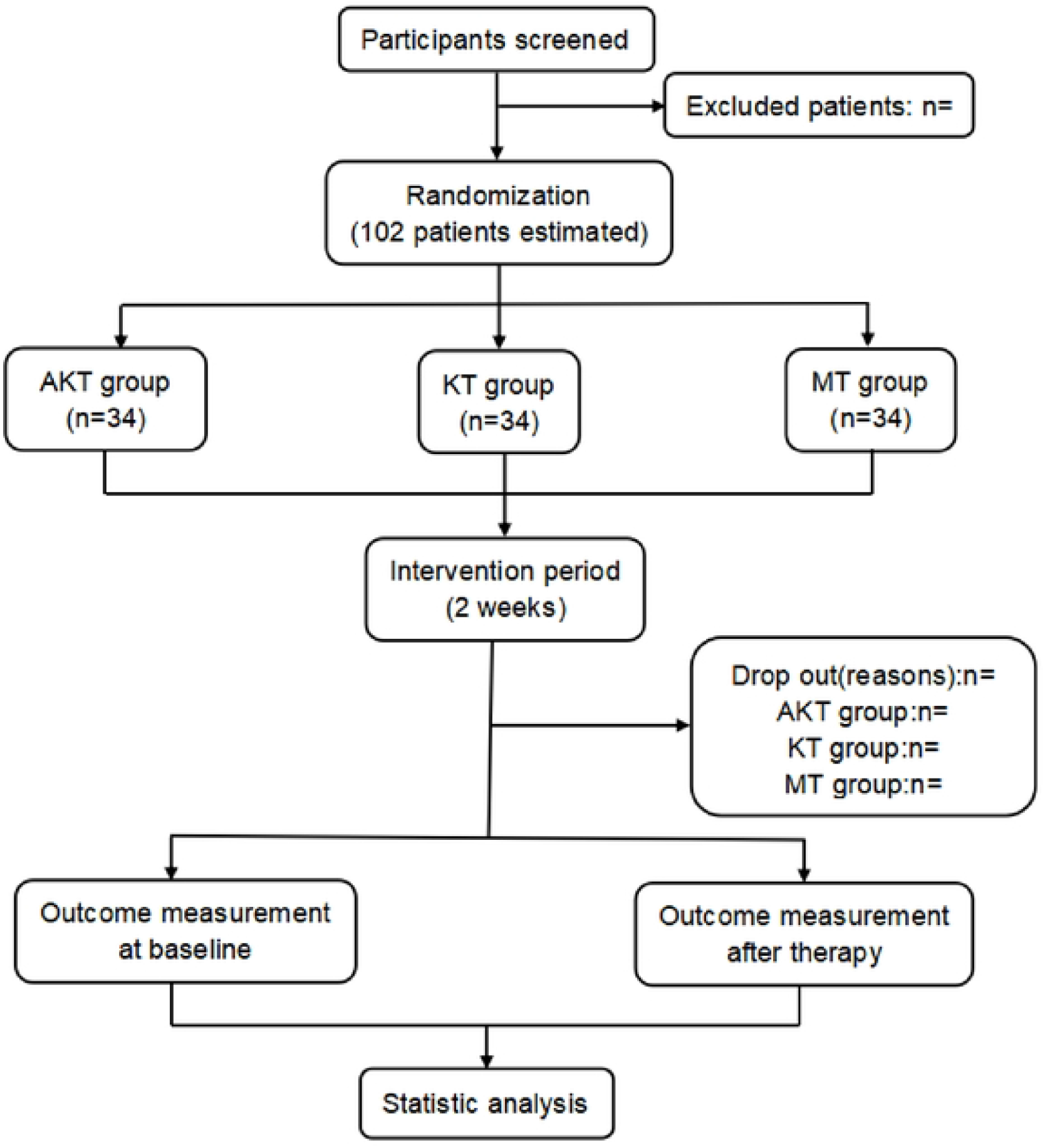
CONSORT flow diagram

### 2.2 Participants

#### 2.2.1 Diagnostic criteria

Patients conform to the diagnostic criteria of the 2016 American College Of Cardiology / American Heart Association Guideline Focused Update on Duration of Dual Antiplatelet Therapy in Patients With Coronary Artery Disease [37] can be recruited, the diagnostic criteria are shown following:

1. The patient had a history of CHD with angina pectoris.
2. Changes in ECG
  ① stable angina pectoris: ECG is almost in the normal range during the remission period. During the attack of angina pectoris, the ST-segment depression dominated by R-wave and T-wave is flat or inverted (ST-segment elevation in the lead of variant angina pectoris), which gradually recovers within a few minutes after the attack.
  ② ST-segment elevation myocardial infarction: The ST-segment arched upward (unidirectional curve) with or without pathological Q and R wave reduction (ST-segment changes are not obvious in posterior wall myocardial infarction) in the early time. The hyperacute phase may be characterized by unusually tall and two asymmetric T waves.
  ③ Non-ST-segment lifting myocardial infarction: the ST-segment depression and T wave inversion present dynamic changes. The T wave inversion gradually deepens and then becomes shallower, and some abnormal Q waves appeared.
3. For patients with myocardial necrosis, the concentrations of serum myocardial markers, such as cTn, myoglobin, CK, and CK-MB are abnormally increased.
4. Coronary angiography shows that the stenosis of the coronary artery is more than 50%.

#### 2.2.2 Inclusion criteria

Patients who fulfill all the following inclusion criteria will be included in this study(**Table 1**).

**Table 1.**
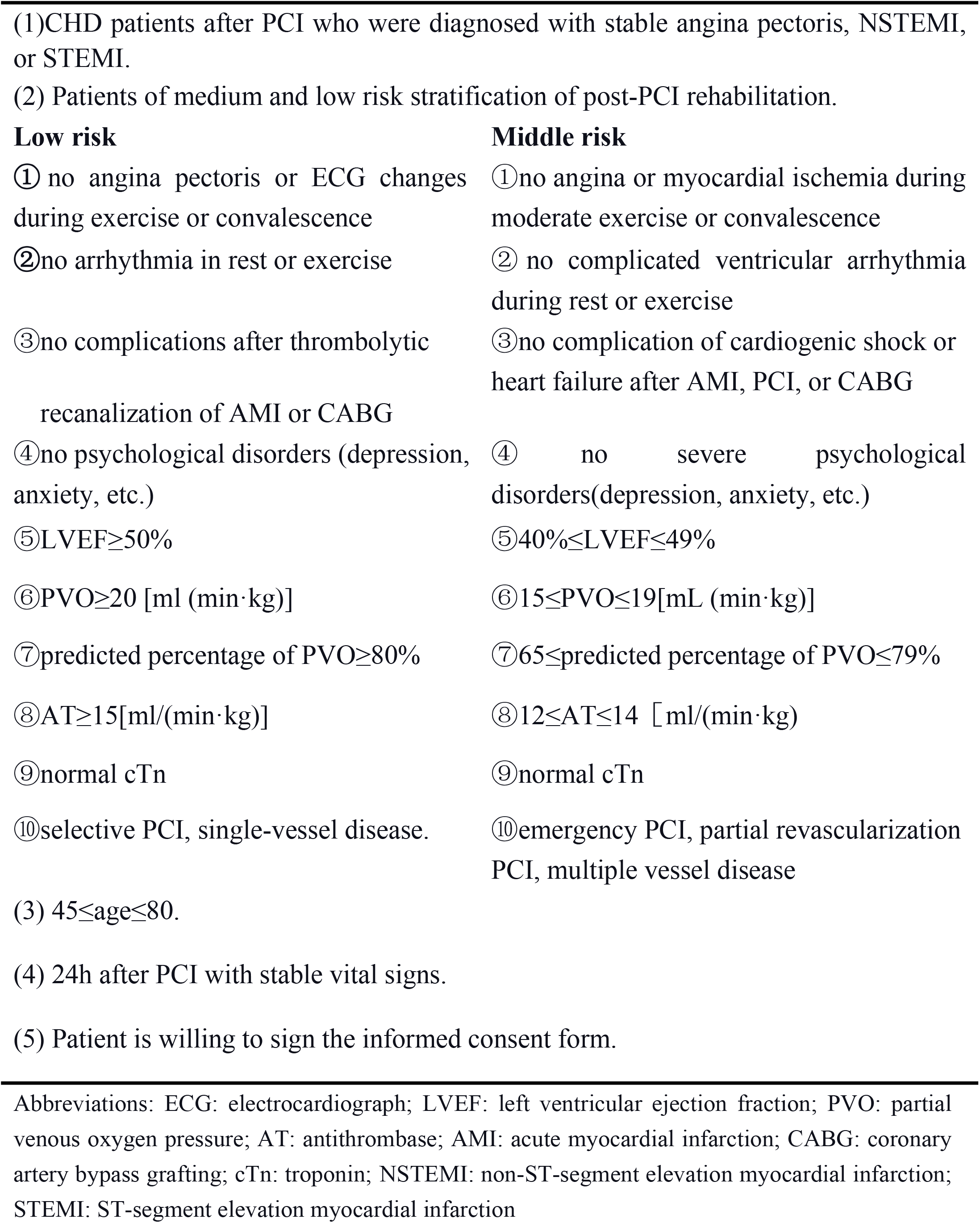
Inclusion criteria.

#### 2.2.3 Exclusion and withdrawal criteria

Patients who meet one of the following criteria will be excluded and allowed or required to withdraw from the trial (**Table 2**). The reasons and exit time will be recorded in standard case report forms (CRF).

**Table 2.** Exclusion and withdrawal criteria

| Exclusion criteria | Withdrawal criteria |
| --- | --- |
| (1) Patients with unstable vital signs. | (1) participants developing a serious disease which is not suitable to continue in the investigator's opinion. |
| (2) Patients with serious complications such as malignancy, heart failure, respiratory failure, shock, etc. | (2) seriously adverse events happen. |
| (3) Patients with cognitive impairment, severe hearing and visual impairment. | (3) participants request to withdraw from the trial. |
| (4) Patients with severe liver, kidney, and immune dysfunction. |  |
| (5) Pregnant women and lactating patients. |  |
| (6) Patients cannot be treated with acupuncture. |  |

#### 2.2.4 Attendance and drop-outs

The attendance of each participant will be checked and recorded in the case report form(CRF). Drop-outs from causes such as house moving, the onset of severe diseases, or other medical complications are unavoidable, all the details will be recorded in the CRF.

#### 2.2.5 Recruitment

Participants will be recruited by posters, doctor’s recommendations, the internet and the WeChat in Chengdu city. If participants meet the study criteria, they will be required to sign an informed consent form prior to the inclusion of the study.

### 2.3 Randomisation and allocation concealment

Included participants will be randomly assigned into 3 groups (AKT group, KT group, and MT group) with a 1:1:1 ratio. The randomization sequence will be created using R3.5.1 by an independent statistician. The information of random numbers, allocation and intervention will be packed in opaque envelopes, and will be concealed to screeners and outcome assessors. Eligible participants will be informed with the allocation results through phone call after opening envelopes in order.

### 2.4 Blinding

The single blind method will be conducted in this study according to the nature of acupuncture and exercise. Participants and researchers are impossible to be blinded to the group assignment. All the outcomes will be measured by the same independent experienced assessors who are blinded to the allocation. After completing the statistical analyses, the group assignment will be revealed by the project manager.

### 2.5 Sample size

According to a previous study [38], the mean difference and standard deviation of ejection fraction (EF), an index of cardiac function, were 52.15 ±1.91, 57.56±2.13 and 59.92±1.93 for CHD patients after PCI, when comparing the effectiveness of AKT compared with KT and MT control. The sample size was calculated using One-Way analysis of variance, F-Tests by the G. Power software. A sample size of 93 participants is required to sufficiently detect a target effect size. Considering a 10% drop-out rate, we intend to enroll a total of 102 participants, with 34 participants in each group.

### 2.6 Intervention

All participants will be asked to treat with MT after the vital signs are stable. We will use CRF to record the intervention progress of participants in the AKT, KT, and MT groups. To monitor the progress, we will collect and analyze the session attendance as well as the self-reported diaries.

1. AKT group: according to the standard treatment of ocular acupuncture approved by the Standardization Administration of China in 2009 [27], the “Upper-Jiao Area” and the “Heart Area” will be selected and penetrated horizontally 2mm outside the orbital margin using the specially-made ocular acupuncture instrument. The ocular acupuncture instrument will be kept embedding until the daily following KT is completed. KT refers to experts consensus of exercise rehabilitation after PCI in patients with CHD published by the Chinese Journal of Interventional Cardiology in 2016 [39]. KT, including warm-up exercises (low levels of aerobic exercise), exercise training(aerobic training, walking training, treadmill training, resistance training, upper limb flexion adduction and abduction exercise, lower extremity knee flexion and extension exercise, and flexibility training), and relaxation training (stretching exercise), will be performed immediately based on the conditions of patients after the acupuncture. It will be manipulated once a day, 5 days a week, and lasting for a total of 2 weeks with a 2-day off during the course. We also will adjust the prescription of exercise according to the patient’s sweating, breathing, pulse and blood pressure.
2. KT group: the KT program will be the same as that in the AKT group.
3. MT group: MT refers to the Chinese Guidelines for PCI published by the Chinese Journal of Cardiology in 2016 [40]. Aspirin enteric-coated tablets, Clopidogrel bisulfate tablets, and the Atorvastatin Calcium Tablets will be applied. Subjects will be asked to take aspirin enteric-coated tablets before (100∼300mg) and after the PCI (100mg, QD, 2 weeks). Clopidogrel bisulfate tablets will be taken at least 6h before surgery (300∼600mg), 2h∼6h before surgery (600mg), and after the PCI (75mg, QD, 2 weeks). Oral Atorvastatin Calcium Tablets will be administered 10mg/d for 2 weeks.

During the study, subjects will also be given antihypertensive and hypoglycemic drugs according to their complications. After the study, follow-up treatment will be conducted under the guidance of a professional cardiologist.

### 2.7 Baseline assessment

The participants will undergo baseline assessment before intervention. A descriptive exploratory questionnaire will be administered to collect information as follows:

1. Demographic and socioeconomic variables (height, weight, gender, age, marital status, education, history, family history, etc.);
2. Clinical variables (temperature, blood pressure, pulse respiration, liver function, kidney function, etc.);
3. General characteristic variables (course of CHD; interventional duration of PCI; the location and extent of coronary artery lesions; complete revascularizations or not; PCI-related complications or not; postoperative medications; rehabilitation plans; rehabilitation beginning time; treatment course, etc.)

### 2.8 Outcome measurements

#### 2.8.1 Primary outcome

The primary outcome is the cardiac function between baseline and 2-week after treatment.

1. 24h Holter Electrocardiogram. The 24-hour Holter electrocardiogram will be used to record the 24h average/lowest/highest heart rate rhythm ST-T changes, heart rate variability (HRV, SDA, HF, LF, LF/HF) and other indicators of the subjects to evaluate the arrhythmias in PCI myocardial ischemia and changes in autonomic nerve function.
2. Color Doppler Echocardiography. The systolic and diastolic functions of the subjects will be evaluated by using doppler echocardiography to record the SV, CO, FS, EF, IVRT, and E/A.
3. Cardiac markers. Mb, cTn, CK, CK-MB α, TNF-α, hs-CRP, and BNP will be detected by venous blood collection. IL-6 levels will be evaluated for ischemic necrosis of heart injury

#### 2.8.2 Secondary outcomes

Aerobic capacity (evaluated by 6min walking test), muscular strength (measured by manual muscle test and isokinetic strength), muscle flexibility (measured by Thomas test), coordination (measured by finger nose test, finger-finger test and heel-knee shin test), balance (measured by Berg Balance Scale), psychological states (measured by self-rating anxiety scale and self-Rating depression scale), and activities of daily living (measured by basic activities of daily living) will be considered as secondary outcomes. The secondary outcomes will be evaluated at baseline and 2-week after treatment.

### 2.9 Follow up

The cardiac function and activities of daily living will be followed up at 1, 3 and 6 months after treatment to observe the long-term efficacy.

### 2.10 Safety monitoring

Adverse events are defined as any unexpected or uncomfortable signs, symptoms, or diseases, including fainting needles, bending needles, stagnation needles, subcutaneous hematoma, skin lesions, muscle soreness, abnormal breathing, and recurrence rate of cardiac events. If any adverse events happen during the entire observation period, all the details will be recorded in the CRF. Data Monitoring Committee (DMC) consisted of statistician, assessor, and experts will discuss and determine the patient should continue or not once an adverse event occurs.

### 2.11 Data collection and management

All data will be collected and managed using the Research Electronic Data Capture (REDCap) system [41]. The data will be entered into the REDCap Database by a dependent researcher, then double-checked by a second researcher. The statistician will review the database to ensure accurate data collection and correct data export for future analyses. All data will either be kept in the Hospital of Chengdu University of Traditional Chinese Medicine and be backed up in different network drives.

### 2.12 Data analysis

The statistical analysis will be carried out using SPSS statistical package (version 23.0). Data will be expressed as mean ± standard deviations. All primary and secondary analyses will be analyzed based on intention-to-treat. Missing data will be filled by the Last Observation Carry Forward rules. Demographic characteristics and other baseline values will be described using descriptive statistics. Baseline characteristics of participants will be compared using the chi-square test for enumeration data and one-way analysis of variance for continuous variables. Differences between the groups will be calculated as mean differences or odds ratios alongside their 95% confidence intervals. Multiple comparisons between the groups will be adjusted according to the Bonferroni correction method. All tests will be two-sided, with the P value less than 0.05 deemed statistically significant.

### 2.13 Patient and public involvement

No patient has been involved in the design, conception and conduction of this trial. Recruitment and data collection began in May 2022, all participants are expected to complete the trial by December 2023.

### 2.14 Ethics and dissemination

This trial will be conducted according to the principles of the Declaration of Helsinki, which has been approved by the Institutional Review Board of Hospital of Chengdu University of Traditional Chinese Medicine in July 2021 (reference number: 2021KL-028). If there is any modification to the protocol which may impact the conduction of this study, the potential benefit of the patient or may affect patient safety.

We will draft a formal amendment to the ethical review committee for approval prior to implementation. This clinical trial is also registered with an identifier (ChiCTR2100048960) at the Chinese clinical trial registry in July 2021. Risks and benefits will be explained clearly to the participants, and they will be given enough time to ask questions and decide whether they will participate in this trial. The authors intend to publish the findings of the study in peer-reviewed journals.

## 3. Discussion

This study aims to conduct a randomized controlled trial (RCT) to confirm the superior efficacy of AKT compared to KT and conventional medical therapy (MT) in post-PCI rehabilitation for patients of CHD, and to further guide the clinical strategy.

The tissue around the eye is closely related to the Zang and Fu organs, and acupuncture around the eye is effective in treating heart and heart diseases. According to the Meridian Theory of the Traditional Chinese Medicine [30,31], the Hand Shaoyin Meridian of Heart, the Hand Shaoyin Divergent Meridian, and the Hand Shaoyin Collateral circulate from the heart and the root of the tongue up to and around the eyes. It is said that the heart controls people’s minds and spirits, once the mind is stable, people would sleep well. Therefore, acupuncture around the eyes was confirmed effective in treating insomnia patients of heart-kidney imbalance type, heart-spleen deficiency type, and deficiency of yin and excess of heat type [42-46]. Moreover, acupuncture on the acupoints of the “Heart Area” and the “Upper Jiao Area” around the eye can significantly reduce the time and frequency of angina pectoris in patients with CHD, as well as the levels of TNF-α and CRP [32]. These findings provide theoretical basis and clinical basis for the use of acupuncture therapy in the rehabilitation of CHD patients after PCI.

We will conduct strict quality control to achieve high reliability and reproducibility. We will consult experts to formulate the Standard Operating Procedure (SOP) of this study. Researchers involved in this study will receive theoretical and practical training of SOP in screening eligible participants, completing the CRF, and assessing outcomes. To improve the baseline consistency of participants, we will strictly follow the inclusion and exclusion criteria for participant enrollment. In addition, researchers and outcome assessors will be separated during the study period. Allocation of participants will be concealed to outcome assessors and data analysts. All researchers will conduct this trial in accordance with SOP. A monthly quality control assessment will be performed to ensure research quality. The DMC will supervise the study.

In this study, we expect findings of trial will provide important insight into application of AKT as a safety and more effective method for CHD patients. Findings of this study are expected to provide an evidence-based decision on the application of AKT and to optimize the post-PCI rehabilitation program by reducing complications after PCI, prolonging postoperative survival time, and improving quality of life for CHD patients after PCI.

## Data Availability

The data underlying the results presented in the study are available from (include the name of the third party and contact information or URL).

## Data Availability

The data and materials are available upon request from the corresponding author.

## Authors’ contributions

Conceptualization: Di Zhang, Huaimin Lu, Song Jin, Hong Guo.

Funding acquisition: Di Zhang.

Investigation: Jiang Ma, Xiaoxiao Liu, Wensheng Xiao, Hongpeng Li.

Methodology: Jiang Ma, Di Zhang.

Project administration: Hongpeng Li.

Supervision: Song Jin, Hong Guo.

Writing–original draft: Jiang Ma.

Writing–review & editing: Di Zhang, Huaimin Lu, Song Jin, Hong Guo, Xiaoxiao

Liu, Wensheng Xiao, Hongpeng Li.

